# Early Photoreceptor Disruption in Emerging Subretinal Drusenoid Deposits Detected by Adaptive Optics Imaging

**DOI:** 10.64898/2026.01.28.26344907

**Authors:** Sujin Hoshi, Xiaolin Wang, Shin Kadomoto, Ruixue Liu, Michael Ip, Srinivas R Sadda, David Sarraf, Yuhua Zhang

**Author notes:** **Corresponding Author:** Yuhua Zhang, PhD, Address: 150 N Orange Grove Boulevard, Pasadena, CA 91103, USA. **Declaration:** This manuscript describes original work that has not been published or submitted elsewhere. **Ethics:** The study followed the tenets of the Declaration of Helsinki, complied with the Health Insurance Portability and Accountability Act of 1996, and was approved by the Institutional Review Boards at the University of California at Los Angeles. Written informed consent was obtained from all participants. **Data Availability:** All data produced in the present work are contained in the manuscript.

## Abstract

**Purpose:** Subretinal drusenoid deposits (SDDs) are a distinct entity in age-related macular degeneration (AMD) and associated with photoreceptor impairment during progression. Their early impact on photoreceptors remains incompletely understood. This study examined photoreceptor reflectivity during the phase when SDDs were not clinically detectable on optical coherence tomography (OCT) using adaptive optics scanning laser ophthalmoscopy (AOSLO).

**Design:** Longitudinal observational study.

**Participants:** Patients with intermediate AMD.

**Methods:** Six eyes of four patients with intermediate AMD and predominantly SDDs underwent multimodal imaging 3-4 times over 3.5 years. Individual SDDs were graded using an OCT-based 3-stage system at each time point. Cross-sectional retinal structure and photoreceptor reflectivity at the location where the new SDDs developed during follow-up were evaluated using OCT and AOSLO.

**Main Outcome Measures:** Photoreceptor reflectivity change prior to and during SDD development.

**Results:** Forty-eight retinal locations where new dot-type SDDs developed during follow-up were identified. AOSLO revealed reduced photoreceptor reflectivity in these regions before OCT demonstrated clinically evident deposits (stage ≥ 1) between the ellipsoid zone and the retinal pigment epithelium at the corresponding sites. The mean time to development of stage 1, stage 2, and stage 3 SDDs was 11.78 ± 5.01, 17.40 ± 6.08, and 18.72 ± 4.08 months, respectively.

**Conclusions:** High-resolution adaptive optics confocal imaging enables detection of photoreceptor optical property alterations at a stage when SDDs are not yet evident on OCT. This finding underscores the exceptional sensitivity of photoreceptors to minimal structural or functional perturbations during SDD formation and defines an early window for potential intervention.

## Introduction

The recognition of subretinal drusenoid deposits (SDDs)^1,2^ as the histologic correlate of reticular pseudodrusen^3-5^ represents an important refinement of the pathogenic framework of age-related macular degeneration (AMD).^6-9^ Classical models focused on drusen accumulating in the sub–retinal pigment epithelium (sub-RPE) space.^10-14^ The finding that SDDs arise instead in the subretinal space between the photoreceptors and RPE revealed a second disease compartment with direct implications for photoreceptor–RPE biology,^2,15^ lipid handling,^2,16^ and outer retinal atrophy.^17^ Because photoreceptors are the essential cells responsible for phototransduction and the earliest stages of visual processing, their contiguous relationship with SDDs may have profound biological and clinical consequences.

Multimodal retinal imaging has greatly advanced characterization of SDD.^6-9,18^ While optical coherence tomography (OCT) provides the defining laminar localization and cross-sectional signature,^3,19^ color fundus photography, near-infrared reflectance, and fundus autofluorescence offer complementary en face spectral contrast.^8^ Clinicopathologic studies demonstrate deflected, shortened, or absent outer and inner segments of photoreceptors overlying SDDs.^2,4,5^ Functional testing reveals localized retinal sensitivity loss.^20-24^ SDDs have been established as a distinct phenotype associated with increased progression to both atrophic and neovascular late AMD.^7,8,25-28^ Moreover, the recent recognition of outer retinal atrophy following SDD regression underscores an unique pathway to the end stage of disease.^17,29^

Adaptive optics (AO) imaging enables cellular level assessment of these processes.^30,31^ Confocal adaptive optics scanning laser ophthalmoscopy (AOSLO) offers sensitive en face visualization of photoreceptor reflectivity, revealing characteristic stage-dependent photoreceptor alterations associated with SDD.^32-34^ Despite these advances, the mechanistic sequence linking SDD formation and early photoreceptor dysfunction remains incompletely defined. Whether SDD initiation stems primarily from RPE dysfunction, or photoreceptor impairment, or a combined disturbance in outer retinal metabolic homeostasis has been an area of active investigation.^2,4,5,35,36^

An important unanswered question concerns the status of photoreceptors before SDDs become detectable using standard clinical diagnostic methods, for which OCT has served as the gold standard.^3,7,37,38^ Existing studies describe photoreceptor alterations only after stage-1 lesion are visible on OCT.^3,18,37^ SDDs in the context of AMD are detected relatively late in life, with currently unknown precursors.^7^ If SDD originate in the subretinal space as a consequence of photoreceptor–RPE dysregulation, perturbations in photoreceptor structure and function, reflected by changes in cellular-level optical properties revealed by adaptive optics imaging, may signal the earliest pathological alterations preceding the accumulation of OCT-detectable material. Identifying such early optical abnormalities in photoreceptors residing in regions destined to develop SDDs could provide critical mechanistic insight into SDD biogenesis. In earlier work, we used AOSLO to characterize SDD morphology and stage-dependent effects on cone photoreceptor reflectivity.^32-34^ During long-term follow-up, some eyes developed new SDDs, providing an unique opportunity to examine AOSLO and OCT features before and after lesion emergence.

Based on the premise that SDDs alter photoreceptor-RPE interactions from their earliest stages, we hypothesized that cone photoreceptor reflectivity would show detectable alterations prior to OCT-visible SDD. In the present study, we leverage longitudinal AOSLO and OCT data to evaluate photoreceptor reflectivity at retinal locations that later developed SDDs, with the goal of clarifying early photoreceptor involvement and refining the mechanistic model of SDD pathogenesis in AMD.

## Methods

The study followed the tenets of the Declaration of Helsinki, complied with the Health Insurance Portability and Accountability Act of 1996, and was approved by the Institutional Review Boards of the University of Alabama at Birmingham and the University of California - Los Angeles. Written informed consent was obtained from all participants.

This was a retrospective study of a subgroup of patients who participated in the adaptive optics imaging of AMD project.^21,39^ The study patient recruitment and multimodal imaging protocol have been reported elsewhere.^21,32-34,39-42^ For the readers’ convenience, we summarize the procedures here.

### Study patients

The participants were recruited from the clinical research registry of the Department of Ophthalmology and Visual Sciences of the University of Alabama at Birmingham. All these subjects were diagnosed with AMD previously, with a best-corrected visual acuity (BCVA) of 20/100 or better, and a refractive error within ±6 diopters spherical and ±3 diopters cylinder. Exclusion criteria included diabetes, history of retinal vascular occlusion, and any historical or examination features of hereditary retinal dystrophy. AMD severity was assessed by a masked, experienced grader, who graded the color fundus photographs taken from the subjects’ eyes using the Age-Related Eye Disease Study 2 (AREDS2) severity scale for AMD. Disease severity ranged from early to advanced (AREDS steps 2-9).^43^ It is worth clarifying that most eyes used in this study showed predominantly SDDs with only a few conventional drusen. Baseline imaging was performed between February 2012 and August 2015. Follow-up imaging was performed four times over a 3.5 year follow-up period.

### Multimodal imaging

In all subjects, color fundus photographs were taken using a fundus camera (Carl Zeiss Meditec Inc., Dublin, CA). En face near-infrared reflectance (NIR; λ = 830 nm), blue reflectance (BR; λ = 488 nm), fundus autofluorescence (FAF; excitation, λ = 488 nm; emission λ > 600 nm), and spectral domain OCT (central macula 15° × 15° to 20° × 20°, 97 - 121 B-scans) were acquired using a Spectralis imaging station (Heidelberg Engineering, Carlsbad, CA).

### AOSLO

was performed to image the macular photoreceptors using a lab developed research instrument that employs a low coherent near-infrared superluminescent diode (λ = 840 nm),^44-47^ through a dilated pupil (with 1.0% tropicamide and 2.5% phenylephrine hydrochloride). The AOSLO images were continuously recorded across the macula (15° × 15° to 20° × 20°). Post-processing corrected the nonlinear distortion caused by the resonant scanner, registered successive frames to enhance the signal-to-noise ratio, and montaged all individual images with cell-to-cell precision using custom and commercial software (Photoshop, Adobe Systems Inc., Mountain View, CA). The AOSLO montage was registered to overlay on the color fundus photograph, NIR, FAF, BR images, and OCT *en face* image using custom software.

### Study protocol for SDD grading

#### SDD grading

SDDs were examined and confirmed on OCT B-scans and on en face color fundus photographs, NIR, and FAF images.^32^ Lesions were graded using the OCT-based 3-stage system at each visit.^3^ For stage 1 SDDs, OCT shows an undulating EZ with accumulation of granular hyperreflective material between the EZ and RPE, while AOSLO reveals focal areas of reduced photoreceptor reflectivity with an undetectable cone mosaic. For stage 2 lesions, OCT demonstrates further inward displacement of the EZ by mounds of subretinal material, and AOSLO shows an expanded area in which photoreceptors are no longer visible. For stage 3 lesions, SDDs interrupt the EZ and extend toward the external limiting membrane (ELM). AOSLO reveals a distinct hyporeflective annular zone with indistinct photoreceptors surrounding a reflective granular center. Outside the hyporeflective annulus, the photoreceptors reappear with variable reflectivity.^32-34^

#### Identifying newly developed dot-form SDDs

Although both dot and ribbon SDDs were present, only dot SDDs were analyzed in this study because the ribbon type was mostly beyond the AOSLO imaging scope. Guided by our previous observations,^32^ we first surveyed follow-up AOSLO images for lesions exhibiting features of newly developed SDDs.

Once a newly developed lesion was identified on AOSLO, corresponding OCT B-scans across this lesion were examined to confirm the presence of SDD. AOSLO cone reflectivity and OCT images from earlier visits at the same retinal locations were then reviewed. On OCT, sites without definite material between the EZ and the RPE were graded as stage 0. To avoid misclassification, B-scan locations were carefully verified on en face images to ensure accurate traversal of the lesion; only lesions with unambiguous OCT identification and clearly aligned B-scans were included, and uncertain cases were excluded to prevent stage underestimation. Cone reflectivity within the lesion zone was assessed visually on AOSLO relative to the surrounding retina. The reflectivity trough-to-edge ratios were measured in the new SDD developed site at each visit, using the function from the software ImageJ (Analyze > Plot Profile, ImageJ1.54f, https://imagej.net/ij/). For each new SDD, the interval between the last visit graded as stage 0 and the first visit at which SDD was detectable on OCT (stage 1–3) was recorded.

#### Measuring local chorioretinal structure in regions with SDD development

To determine whether retinal areas that developed SDDs exhibited greater chorioretinal degeneration, photoreceptor outer nuclear layer (ONL) including the Helen’s fiber layer (HFL) thickness, photoreceptor length, and choroidal thickness were measured at the location where new SDDs appeared and at the same location before their appearance. Measurements were obtained from OCT using the Spectralis software (Heidelberg Eye Explorer, version 1.7.0.0; Heidelberg Engineering). The same parameters were also measured in a control area within 100 μm of the lesion site. The control areas maintained visible cone reflectivity and a contiguous photoreceptor mosaic on AOSLO, with no discernible lesions on OCT throughout the follow-up period.

Following the method of Spaide,^17,40^ choroidal thickness was measured from the outer border of the RPE-Bruch membrane complex to the choroidal–scleral junction. Photoreceptor length was measured from the inner boundary of the outer plexiform layer (OPL) to the midpoint of the RPE–Bruch membrane band. When we measured photoreceptor length at the selected sites, we carefully examined the corresponding OCT B-scans to ensure that measurements were performed where the SDD was present and the RPE band could be clearly delineated. ONL+ HFL thickness was measured from the inner boundary of the OPL to the midpoint of the ELM band.

#### Statistical analysis

BCVA of the participants were converted to logarithm of the minimum angle of resolution (logMAR). Subject age, choroidal thickness, ONL+HFL thickness, and photoreceptor length were summarized as mean ± standard deviation. Choroidal thickness, ONL+HFL thickness, and photoreceptor length at retinal locations where new SDDs developed and the corresponding control regions were compared using paired t-tests (MATLAB R2023a; MathWorks, Natick, MA). A p-value < 0.05 was considered statistically significant. Differences in choroidal and retinal thickness between paired regions were visualized using box plots. Linear mixed-effects models with post-hoc analyses were used to further evaluate the measured structural parameters.

## Results

Six eyes from 4 participants with intermediate-stage AMD were included in the study (**Table 1**).

**Table 1.** Study subjects.

| Subject | Age | Gender | Eye | AREDS grade | LogMAR |
| --- | --- | --- | --- | --- | --- |
| 1 | Middle 80's | Female | OD | 7 | -0.1 |
|  |  |  | OS | 5 | 0.4 |
| 2 | Late 70's | Female | OD | 6 | 0.4 |
|  |  |  | OS | 7 | 0.5 |
| 3 | Middle 70's | Male | OS | 6 | 0.1 |
| 4 | Middle 70's | Female | OS | 6 | 0.1 |

Longitudinal AOSLO disclosed the development and progression of SDDs from stage 0 to stage 3 (**Figure 1**), accompanied by characteristic stage-dependent alterations in photoreceptor reflectivity.^32^ Retina affected by stage 1 lesions exhibited focal areas of hyporeflective cones (**green arrows, Figure 1**). Stage 2 lesions were associated with enlarged hyporeflective regions encompassing a greater number of disrupted photoreceptors with further reduced reflectivity (**magenta arrows, Figure 1**), consistent with outer-segment perturbation. Stage 3 lesions demonstrated a distinctive hyporeflective annulus composed of deflected or degenerate photoreceptors surrounding a central hyperreflective core containing SDD material and degenerated photoreceptors (**red arrows, Figure 1**).

**Figure 1.**
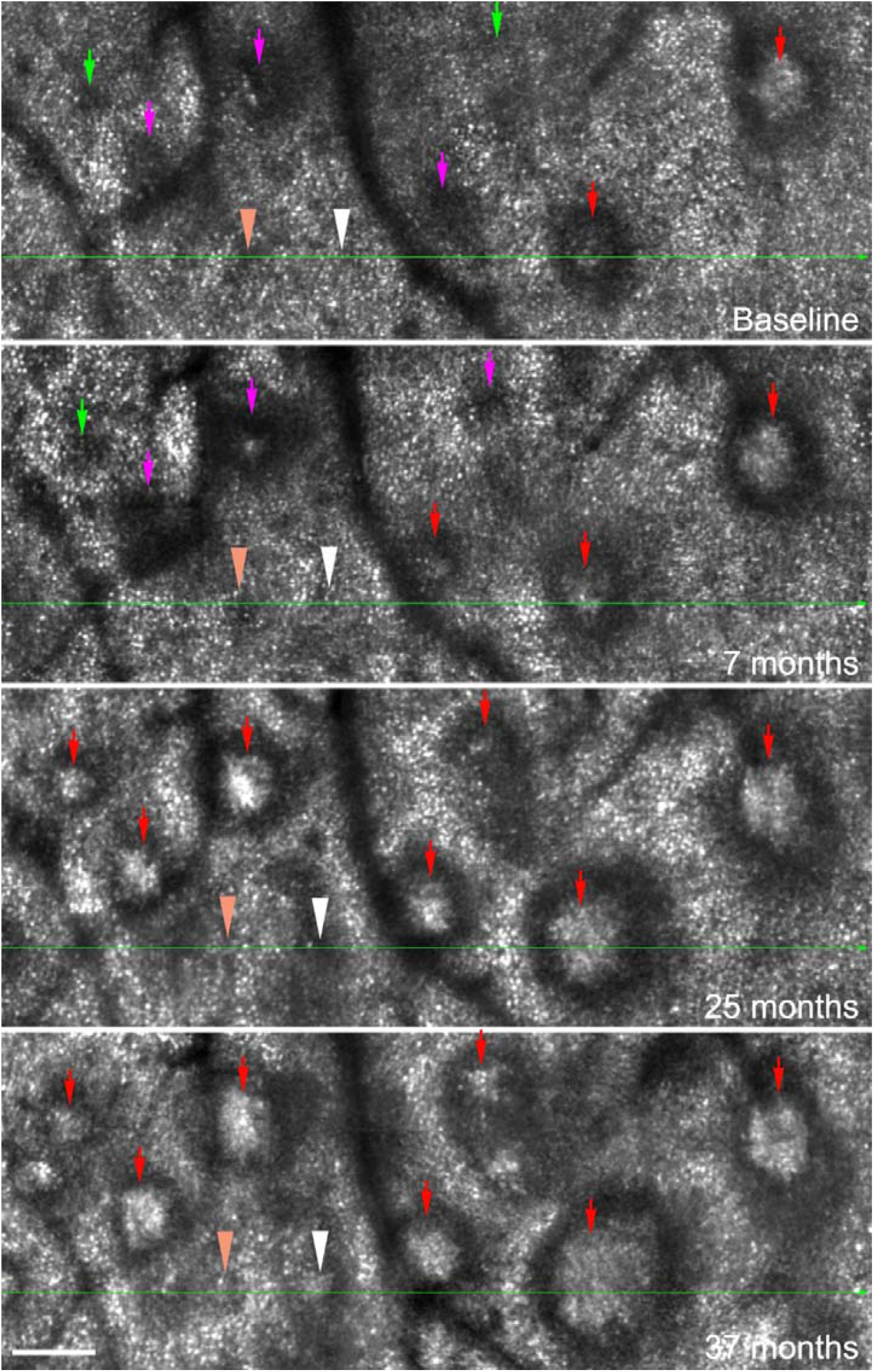
Longitudinal study showing subretinal drusenoid deposits (SDDs) development and progression on AOSLO. The green lines indicate the OCT B-scan position in Figure 2. The orange and white arrowheads show the SDDs correspond to that in Figure 2. On baseline and 7 months, there are no SDDs appearing on OCT, however, the retinal reflectivity on these two areas have already reduced for the orange arrowhead on baseline and for the white arrowhead on 7 months on AOSLO. They show stage 2 SDD (orange arrowhead) and stage 3 SDD (white arrowhead) in 37 months on OCT. The other arrows show SDD progression, with green arrows indicate stage 1 SDDs, magenta arrows indicate stage 2, and red arrows indicate stage 3. The scale bar is 100 µm.

Newly developed SDDs were identifiable on AOSLO before becoming apparent on OCT (**orange and white arrowheads, Figures 1 and 2**). At the visit immediately preceding the first detection of new lesions during follow-up (**7 month visit, Figures 1 and 2**), OCT showed no detectable material between the ellipsoid zone and the RPE (stage 0), whereas AOSLO revealed localized cone hyporeflectivity at the corresponding sites. By 25 months, stage 1 SDDs became apparent on OCT and progressed to stage 3 by 37 months (**Figures 1 and 2**).

**Figure 2.**
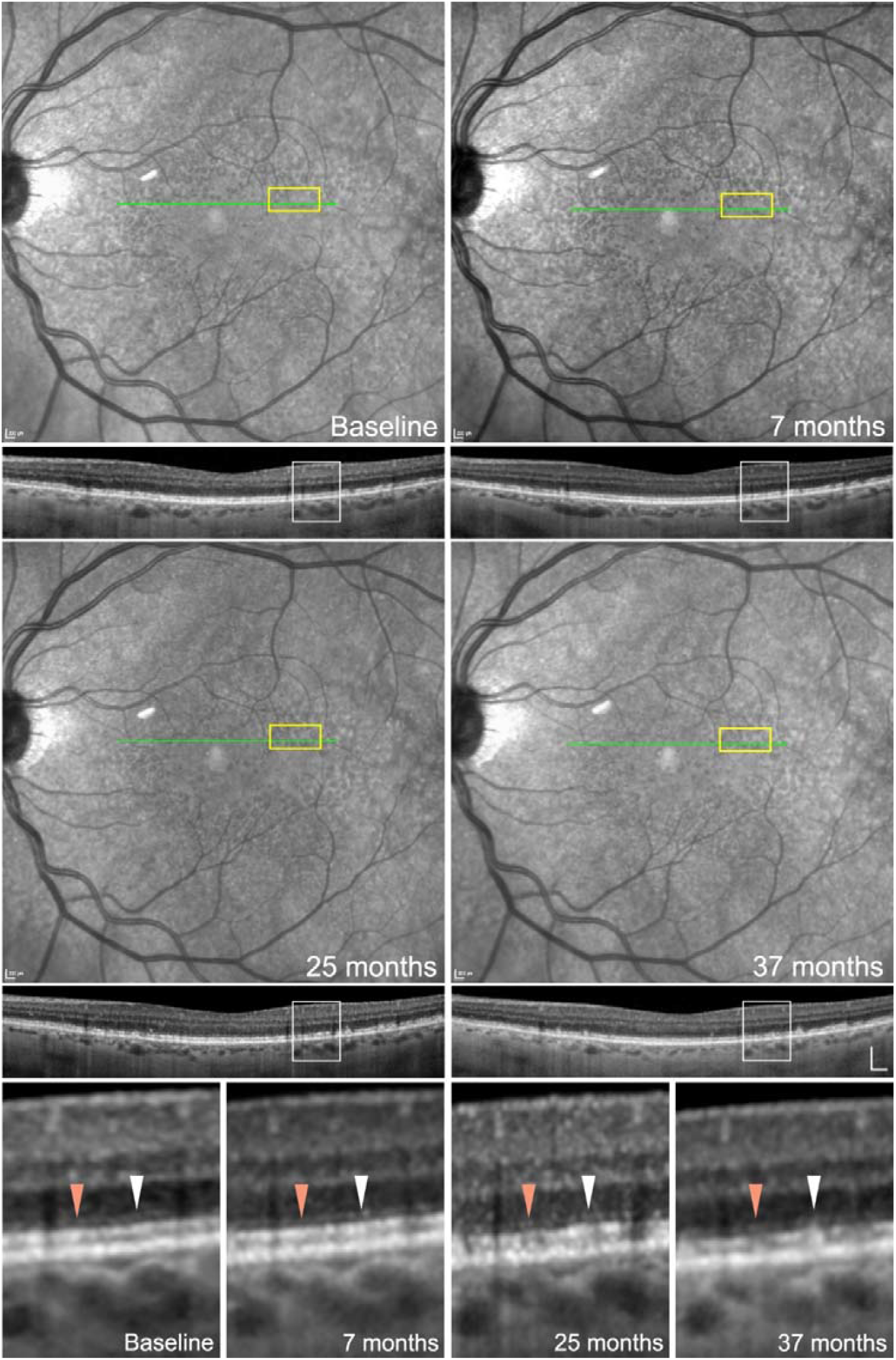
Longitudinal study showing the development of subretinal drusenoid deposits (SDDs) over time on infrared reflectance (IR) and OCT images. The bottom row shows the zoomed-in region outlined by the white rectangles on the OCT B-scans. The green lines on the IR images show the locations of the corresponding OCT B-scan below, while the yellow rectangle highlights the position of the corresponding AOSLO images in Figure 1. Orange and white arrowheads highlight regions of interest where an SDD lesion evolves from no evidence (Stage 0) to stage 2 (orange) and stage 3 (white) on OCT. The scale bar on OCT is 200 µm.

Across these eyes, 48 retinal locations were identified where new dot-type SDDs subsequently developed, including 10 initially appearing as stage 1, 9 as stage 2, and 29 as stage 3. Prior to OCT-detectable SDD formation, these locations demonstrated reduced photoreceptor reflectivity on AOSLO, quantified by the trough-to-edge reflectivity ratio of the lesion region. For this cohort, at the visit immediately preceding OCT detection, the ratio was 0.77 ± 0.11. **Figure 3** and **Figure 4** show the progression images of SDD from stage 0 (no OCT evidence) to stages 1-3, demonstrating that localized reductions in photoreceptor reflectivity well preceded OCT-detectable SDD formation.

**Figure 3.**
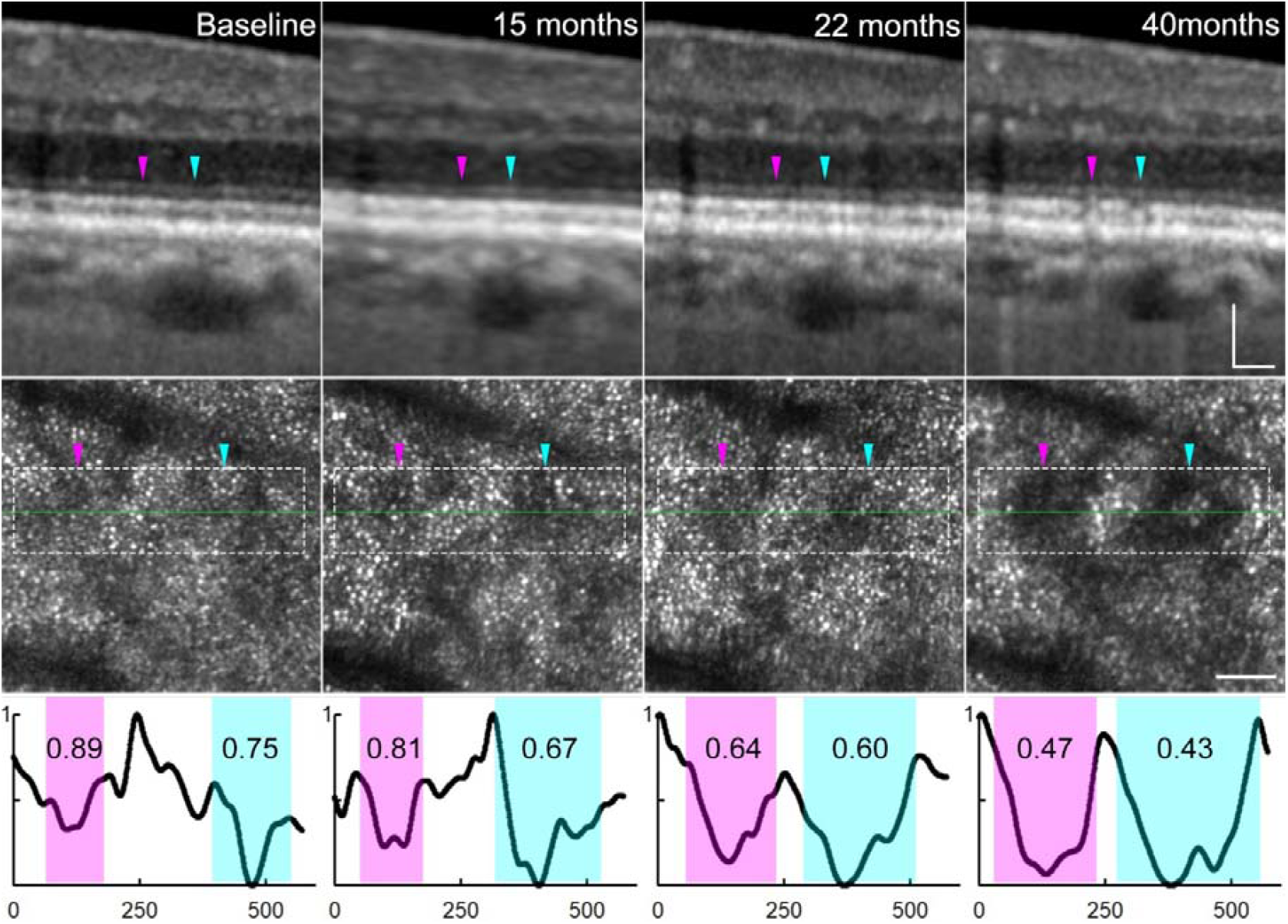
Longitudinal study showing subretinal drusenoid deposit (SDD) development. The green lines on AOSLO images show the position of the corresponding OCT B-scan above. There are no SDD appearing at baseline, 15 months, and 22 months according to the OCT, though stage 2 SDD are evident at 40 months. The locations of the future SDDs exhibit reduced reflectivity compared with the surrounding regions on AOSLO before the SDDs become visible on OCT. The reflectivity profiles (bottom row) from these regions (within the dash-line rectangles) demonstrate reduced reflectivity. The trough-to-edge reflectivity ratios are 0.89, 0.81, 0.64, 0.47 for baseline, 15 months, 22 months, and 40 months, respectively, for the leftmost SDD. They are 0.75, 0.67, 0.60, and 0.43, respectively, for the rightmost SDD region. Magenta and cyan arrowheads and squares indicate the SDD locations. The profile plots were normalized, and the horizontal coordinate unit is in pixels. The scale bars are 100 µm for OCT and 50 µm for AOSLO.

**Figure 4.**
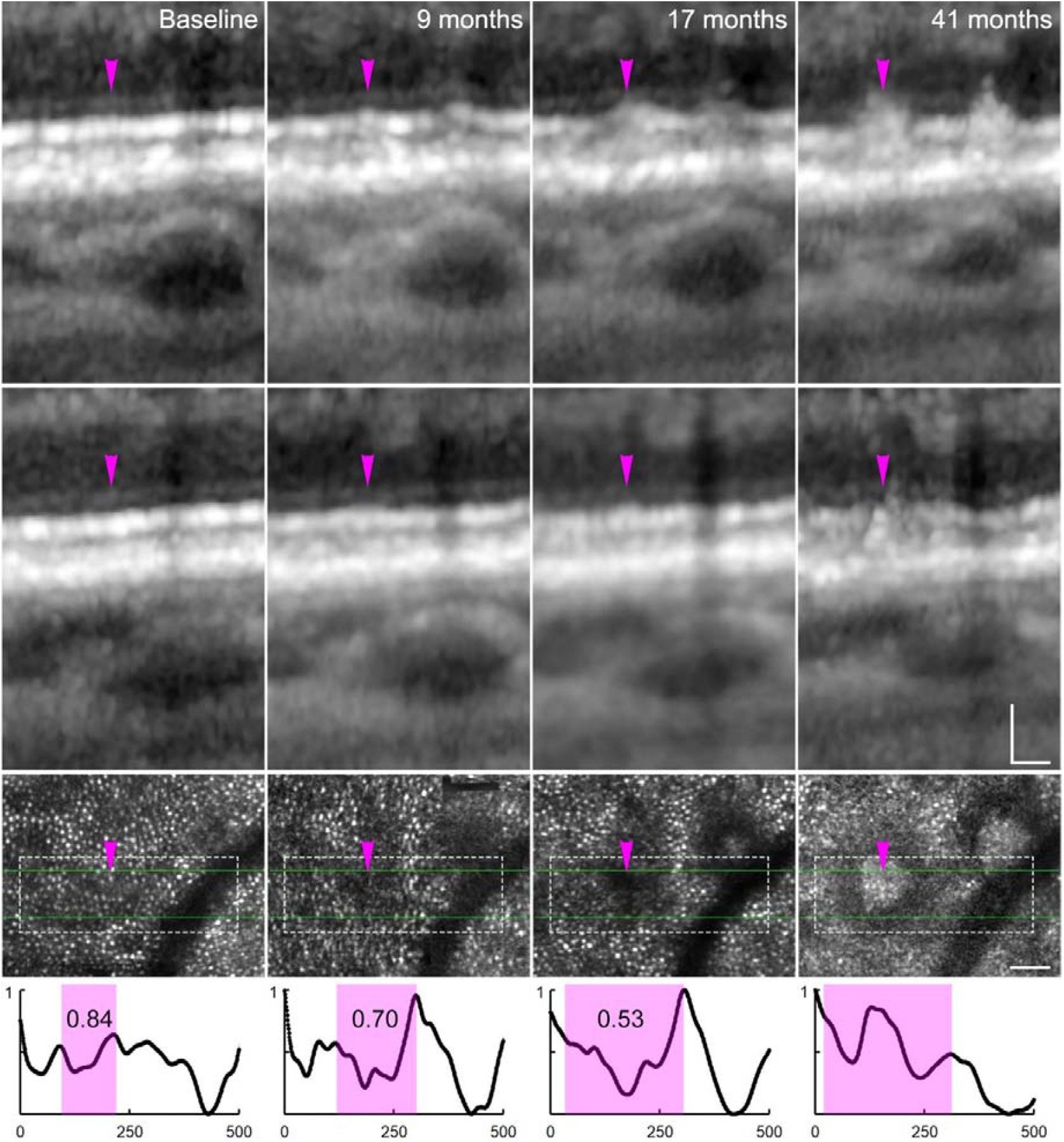
Longitudinal study showing subretinal drusenoid deposits (SDD) progression. The green lines on the AOSLO images show the upper and lower OCT B-scan positions. There are no SDD at either B-scan location at baseline or 9 months by OCT. At the upper B-scan location, a Stage 1 SDD appears at 17 months and progresses to a stage 3 SDD by 41 months. At the lower B-scan location, no SDD is evident at baseline through 17 months but a stage 2 SDD is present by 41 months. It should be noted that both B-scans actually likely pass through the same SDD lesion, but transect the lesion at different points. Regardless, reduced reflectivity can be noted on AOSLO before the SDD is captured with OCT. The reflectivity profiles (bottom row) from these areas (within the dash-line rectangles) confirm this reduced reflectivity. The trough-to-edge reflectivity ratios are 0.84, 0.70, 0.53 for baseline, 9 months, 17 months respectively. Magenta arrowheads and squares indicate the SDD areas. The profile plots were normalized, and the horizontal coordinate unit is in pixels. The scale bars are 50 µm on both OCT and AOSLO.

Of note, many hyporeflective regions on AOSLO later developed SDDs but were not captured by OCT B-scans; these regions were not included in analysis. The mean interval from absence of SDD to OCT-detectable SDD was 16.88 ± 5.80 months. The mean time to development of stage 1, stage 2, and stage 3 SDDs was 11.78 ± 5.01, 17.40 ± 6.08, and 18.72 ± 4.08 months, respectively.

Longitudinal OCT structural analysis (**Table 2**) revealed differential chorioretinal structure changes associated with the development of SDDs (**Figure 5**). At the time immediately preceding the development of SDDs, photoreceptor length, ONL+HFL thickness, and choroidal thickness did not differ significantly between sites where SDDs developed subsequently and the corresponding control areas (all p > 0.44). At follow-up, however, photoreceptor length was significantly reduced at lesion developed sites compared with controls (84.42 ± 28.25 µm vs 130.10 ± 18.60 µm, p < 0.0001), with a significant group-by-time interaction indicating a pronounced decline exclusively in regions where SDDs developed. ONL+HFL thickness similarly showed a significant reduction at lesion sites at follow-up (56.63 ± 14.39 µm vs 61.58 ± 14.12 µm, p = 0.0117), again with a significant interaction reflecting selective thinning associated with SDD appearance. Choroidal thickness decreased significantly over time in both lesion and control regions (both p < 0.003), indicating a generalized thinning over time in this cohort; however, the rate of decline was slightly greater at lesion sites, suggesting some element of more localized choroidal involvement in regions that subsequently developed SDDs.

**Table 2.** Chorioretinal structure changes associated with SDD development (Mean ± SD)

| Thickness ($\mu\text{m}$) | Sites with newly developed SDD | | Control regions<sup>†</sup> | |
| --- | --- | --- | --- | --- |
| | Last time stage 0 | First time stage $\geq 1$ | Last time stage 0 | First time stage $\geq 1$ |
| Photoreceptor | $130.02 \pm 16.63$ | $84.42 \pm 28.25$ | $131.77 \pm 15.79$ | $130.10 \pm 18.60$ |
| ONL+ HFL | $61.02 \pm 13.05$ | $56.63 \pm 14.39$ | $62.52 \pm 13.61$ | $61.58 \pm 14.12$ |
| Choroid | $135.10 \pm 38.26$ | $124.62 \pm 39.23$ | $134.10 \pm 37.92$ | $124.56 \pm 38.02$ |

<sup>†</sup> The parameters were measured in the corresponding control regions at the visits immediately before and after SDD became detectable on OCT.

**Figure 5.**
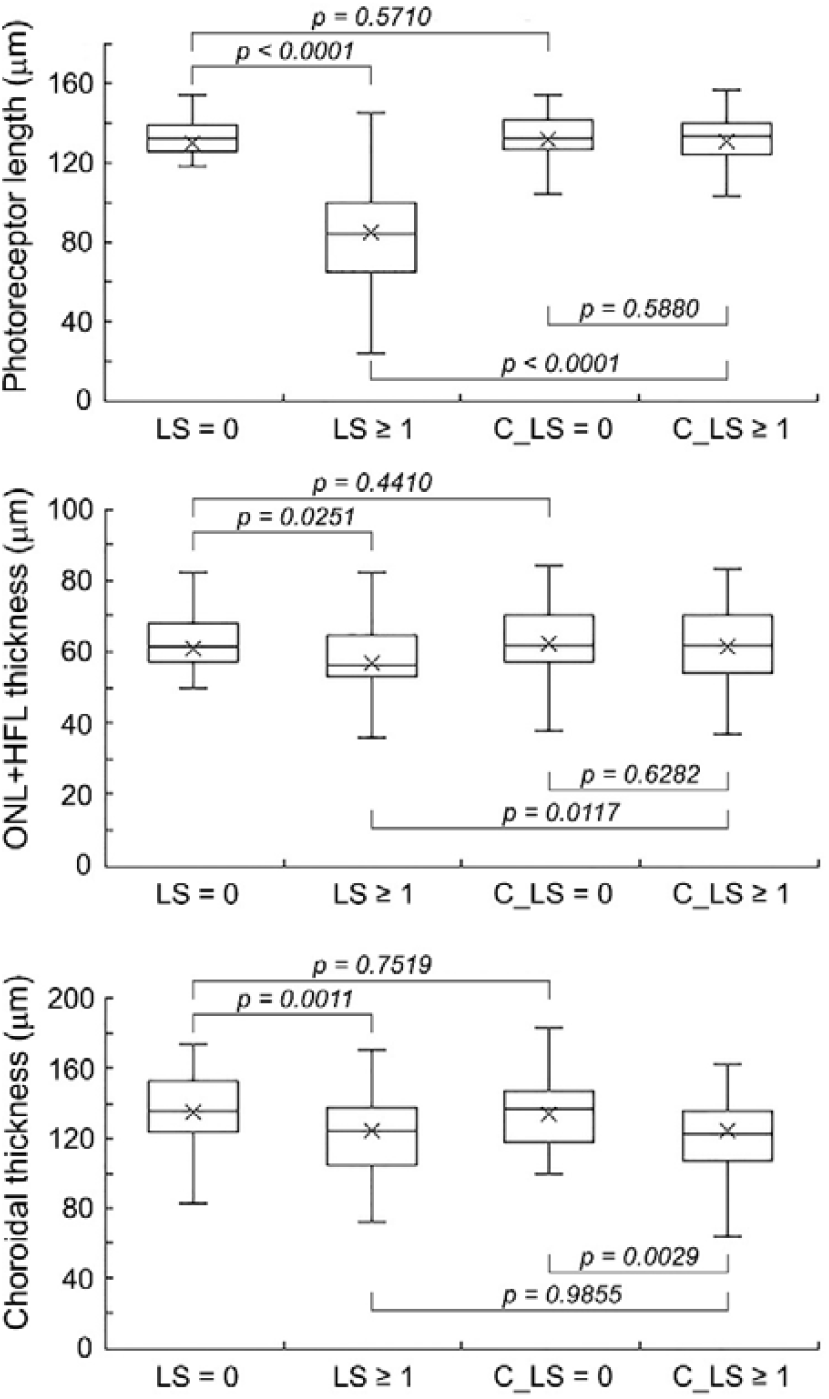
Chorioretinal structural changes associated with SDD development. Photoreceptor length, ONL+HFL thickness, and choroidal thickness were measured at retinal sites where new SDD developed during follow-up and at corresponding control regions. LS = 0 denotes lesion sites at the visit immediately before SDD became detectable on OCT (stage 0); LS ≥ 1 denotes lesion sites at the visit when SDD became detectable on OCT (stage ≥ 1). C_LS = 0 denotes the corresponding control regions at the visit before SDD detection, and C_LS ≥ 1 denotes the corresponding control regions at the visit when SDD was detected on OCT. Before SDD became detectable on OCT, photoreceptor length, ONL+HFL thickness, and choroidal thickness in regions that later developed new lesions did not differ significantly from those measured in corresponding control regions. After lesions became visible on OCT, all three parameters showed significant reductions compared with measurements obtained before SDD detection. In control regions, photoreceptor length and ONL+HFL thickness did not change significantly over time, whereas choroidal thickness showed a significant decrease before and after SDD detected on OCT. In lesion regions, photoreceptor length and ONL+HFL thickness were significantly reduced compared with control regions after SDD detection. In contrast, choroidal thickness did not differ significantly between lesion and control regions after SDD became detectable on OCT.

## Discussion

The contiguous relationship of SDDs with photoreceptor outer segments has important implications for both the optical properties and cellular integrity of photoreceptors. Using confocal AOSLO, we observed a localized reduction in photoreceptor reflectivity that preceded the capture of SDD with OCT. This finding indicates that ultrastructural or functional alterations of photoreceptors, or impaired RPE–photoreceptor interaction, emerges prior to conventional structural detectability. Our study highlights the sensitivity of AOSLO for detecting subclinical photoreceptor alterations and provides mechanistic insight into SDD biogenesis.

A central finding of this study is the diminished cone photoreceptor reflectivity in retinal regions that subsequently developed SDDs. Several biological mechanisms may account for these early optical changes. Clinicopathologic studies have demonstrated that SDDs exert direct impact on overlying photoreceptors, including outer segment deflection, shortening, and loss, as well as progressive inner segment degeneration.^2,4,5^ Disruption of photoreceptor–RPE interactions by SDDs may impair outer segment disc shedding, disrupt morphogenesis, or lead to incomplete renewal, thereby compromising photoreceptor waveguiding efficiency and resulting in reduced reflectivity.^6,7^ RPE dysfunction can contribute, as the RPE governs outer-segment phagocytosis, lipid recycling,^2,4^ and maintenance of photoreceptor orientation.^7^ Additionally, choroidal circulatory insufficiency may produce outer-segment hypoxia, triggering photoreceptor stress and early optical changes.^6,7^ Meanwhile, It is also possible that SDD formation is promoted by photoreceptor stress or dysfunction, raising the possibility that photoreceptors act as upstream contributors to lesion development rather than passive victims.^35^

Although OCT is the clinical gold standard for staging SDDs, distinguishing stage 1 lesions from interdigitation zone (IZ) irregularities can be challenging, particularly in early AMD where IZ visibility is reduced.^48^ In contrast, confocal AOSLO detects cone photoreceptor reflectivity perturbations at an earlier stage. Prior studies have shown that stage 1 SDDs are associated with mild photoreceptor reflectivity loss, progressing to a pronounced hyporeflective annulus in stage 3.^32-34^ The present study extends these observations by demonstrating that reflectivity abnormalities are present even before stage 1 SDDs are captured with OCT. The ability of AOSLO to detect these early perturbations underscores its value in serving as a sensitive technique for identifying disease initiation. Given the lack of available interventions for SDD and early AMD, early identification of functional and structural deficits may facilitate developing novel treatment targeting lesion pathogenesis.

Our observations highlight the remarkable sensitivity of the photoreceptor–RPE complex to even minimal structural or functional perturbations. Clinicopathologic studies have shown that very small SDDs, on the order of ∼4 μm, are associated with deformation of photoreceptor outer segments.^4^ Such physical contact and alteration may produce immediate, localized disruption of outer-segment organization, and compromise waveguiding efficiency. These early changes, however, are difficult to detect with current standard clinical imaging. As shown in this study, at retinal locations that later developed SDDs, photoreceptor reflectivity was already reduced by approximately 23% (**Figure 5**), whereas OCT-derived chorioretinal metrics, including photoreceptor length, ONL+HFL thickness, and choroidal thickness, were indistinguishable from regions that did not subsequently develop lesions. After SDD formation, all these parameters showed significant reductions compared with the visit immediately preceding lesion appearance (**Table 2**).

Notably, early SDD formation was not accompanied by detectable changes in RPE optical properties, such as choroidal hypertransmission, a feature commonly observed in cuticular drusen.^49-51^ This observation is corroborated by clinicopathologic studies showing that the RPE remains largely intact during much of the disease course.^4^ The absence of early OCT-detectable RPE abnormalities, together with preserved RPE cellular structure on histology, raises important questions regarding the timing and nature of the initial pathogenic events that trigger SDD development. Chen and colleagues have proposed that a molecular blockade at the RPE, such as impaired binding or uptake of recycled lipids, could lead to material accumulation and seed subsequent deposition that grows upward toward the external limiting membrane, suggesting a RPE dysfunction-initiated process.^4^ Meanwhile, they also acknowledged that an alternative mechanism with initial disturbance at the photoreceptor level was also theoretically possible.^4^ Although these mechanistic hypotheses require rigorous testing and are beyond the scope of the current study, disruption of photoreceptor–RPE exchange would be expected to adversely affect photoreceptor function. Consistent with this concept, AOSLO revealed reduced photoreceptor reflectivity at sites that later developed SDDs, detectable 11.78 -18.72 months before clinically overt lesions, thereby defining a valuable early window for potential intervention.

A major strength of this study is the combined use of high-resolution AOSLO and OCT to longitudinally track many individual SDDs over an extended follow-up period. By using newly developed, definitively identified SDDs as spatial references, we were able to precisely track the same retinal locations prior to the appearance of clinically overt lesions, enabling accurate characterization of early pathological changes. This study also has limitations. AOSLO primarily visualizes cone photoreceptors, however rod dysfunction is thought to predominate in SDD, and ribbon-type SDDs which tend to distribute in more peripheral locations^52^ were not studied. In addition, the retrospective design and relatively small sample size may limit generalizability. Nonetheless, the integration of high-resolution AOSLO with multimodal imaging provides a uniquely sensitive and definitive assessment of the earliest optical or functional alterations associated with SDD formation.

In conclusion, AOSLO revealed that localized perturbations in photoreceptor reflectivity preceded the formation of OCT-detectable SDDs, supporting a model in which early alterations in photoreceptor outer-segment or RPE function occur at the onset of lesion development. These findings strengthen the hypothesis that SDD formation reflects early dysfunction within the photoreceptor–RPE–choriocapillaris complex, potentially involving impaired lipid recycling, diminished RPE support, or early choroidal insufficiency, before substantial subretinal material accumulates. High-resolution, in vivo assessment of photoreceptor and RPE function will be critical to define the sequence of pathogenic events and to guide the development of interventions targeting the earliest, potentially reversible stages of SDD pathophysiology.

## Data Availability

All data produced in the present work are contained in the manuscript

## Notes

**Funding:** This project was supported by research grants from the National Institute of Health (R01EY024378, R01EY034218, R01EY032994), W. M. Keck Foundation, Carl Marshall Reeves & Mildred Almen Reeves Foundation, Research to Prevent Blindness/Dr. H. James and Carole Free Catalyst Award for Innovative Research Approaches for AMD.

**Conflicts of Interest: Sujin Hoshi, Xiaolin Wang, Shin Kadomoto, Ruixue Liu, Yuhua Zhang**, None. **Michael Ip**, <u>Consultant (C)</u>: Alimera, Allergan, Amgen, Apellis, Astellas, Boehringer Ingelheim, Clearside Biomedical, Genentech, Inc., Novartis, Regeneron Pharmaceuticals, Inc., Zeiss. Recipient (R): Boehringer Ingelheim, 4DMT, Apellis, Astellas, Biogen, Genentech, Lineage Cell Therapeutics, ONL Therapeutics, Regeneron Pharmaceuticals, Inc., and Regenexbio. **David Sarraf**, <u>Consultant (C)</u>: Amgen, Astellas, Eyepoint, Optovue/Visionix. Recipient (R): Amgen, iCARE/Eidon, Optovue/Visionix. **SriniVas R Sadda**, <u>Consultant (C)</u>: Roche/Genentech, Regeneron, Allergan/Abbvie, Novartis, Amgen, Alnylam, Alkeus, Neurotech, 4DMT, Alexion, Nanoscope, Biogen, Samsung Bioepis, Apellis, Astellas, ONL Therapeutics, Optos, Oxurion, Pfizer, Boerhinger Ingelheim, Surrozen, ArrowheadPharma, Eyestem, Topcon, Notal, Heidelberg Engineering, iCare. Recipient (R): Topcon Medical Systems Inc. Heidelberg Engineering, Nidek Incorporated, Novartis Pharma AG; Roche. Financial Support (F): Topcon, Carl Zeiss Meditec, Heidelberg Engineering, Optos Inc., Nidek, iCare/Centervue, Intalight

### Competing Interest Statement

Sujin Hoshi, Xiaolin Wang, Shin Kadomoto, Ruixue Liu, Yuhua Zhang, None
Michael Ip, Consultant (C): Alimera, Allergan, Amgen, Apellis, Astellas, Boehringer Ingelheim, Clearside Biomedical, Genentech, Inc., Novartis, Regeneron Pharmaceuticals, Inc., Zeiss. Recipient (R): Boehringer Ingelheim, 4DMT, Apellis, Astellas, Biogen, Genentech, Lineage Cell Therapeutics, ONL Therapeutics, Regeneron Pharmaceuticals, Inc., and Regenexbio.
SriniVas R Sadda, Consultant (C): Roche/Genentech, Regeneron, Allergan/Abbvie, Novartis, Amgen, Alnylam, Alkeus, Neurotech, 4DMT, Alexion, Nanoscope, Biogen, Samsung Bioepis, Apellis, Astellas, ONL Therapeutics, Optos, Oxurion, Pfizer, Boerhinger Ingelheim, Surrozen, ArrowheadPharma, Eyestem, Topcon, Notal, Heidelberg Engineering, iCare. Recipient (R): Topcon Medical Systems Inc. Heidelberg Engineering, Nidek Incorporated, Novartis Pharma AG; Roche. Financial Support (F): Topcon, Carl Zeiss Meditec, Heidelberg Engineering, Optos Inc., Nidek, iCare/Centervue, Intalight
David Sarraf, Consultant (C): Amgen, Astellas, Eyepoint, Optovue/Visionix. Recipient (R): Amgen, iCARE/Eidon, Optovue/Visionix.

### Funding Statement

This project was supported by research grants from the National Institute of Health (R01EY024378, R01EY034218, R01EY032994), W. M. Keck Foundation, Carl Marshall Reeves & Mildred Almen Reeves Foundation, Research to Prevent Blindness/Dr. H. James and Carole Free Catalyst Award for Innovative Research Approaches for AMD.

## References

1. Rudolf M, Malek G, Messinger JD, Clark ME, Wang L, Curcio CA. Sub-retinal drusenoid deposits in human retina: Organization and composition. Exp Eye Res. 2008;87(5):402–408.

2. Curcio CA, Messinger JD, Sloan KR, McGwin G, Medeiros NE, Spaide RF. Subretinal drusenoid deposits in non-neovascular age-related macular degeneration: Morphology, prevalence, topography, and biogenesis model. Retina. 2013;33(2):265–276.

3. Zweifel SA, Spaide RF, Curcio CA, Malek G, Imamura Y. Reticular pseudodrusen are subretinal drusenoid deposits. Ophthalmology. 2010;117(2):303-312.e301.

4. Chen L, Messinger JD, Zhang Y, Spaide RF, Freund KB, Curcio CA. Subretinal drusenoid deposit in age-related macular degeneration: Histologic insights into initiation, progression to atrophy, and imaging. Retina. 2020;40(4):618–631.

5. Greferath U, Guymer RH, Vessey KA, Brassington K, Fletcher EL. Correlation of histologic features with in vivo imaging of reticular pseudodrusen. Ophthalmology. 2016;123(6):1320–1331.

6. Sivaprasad S, Bird A, Nitiahpapand R, et al. Perspectives on reticular pseudodrusen in age-related macular degeneration. Surv Ophthalmol. 2016;61(5):521–537.

7. Spaide RF, Ooto S, Curcio CA. Subretinal drusenoid deposits aka pseudodrusen. Surv Ophthalmol. 2018;63(6):782–815.

8. Wu Z, Fletcher EL, Kumar H, Greferath U, Guymer RH. Reticular pseudodrusen: A critical phenotype in age-related macular degeneration. Prog Retin Eye Res. 2022;88:101017.

9. Smith RT, Olsen TW, Chong V, et al. Subretinal drusenoid deposits, age-related macular degeneration, and cardiovascular disease. Asia Pac J Ophthalmol (Phila). 2024:100036.

10. Hageman GS, Luthert PJ, Chong NHV, Johnson LV, Anderson DH, Mullins RF. An integrated hypothesis that considers drusen as biomarkers of immune-mediated processes at the rpe-bruch’s membrane interface in aging and age-related macular degeneration. Prog Retin Eye Res. 2001;20(6):705–732.

11. Spaide RF, Curcio CA. Drusen characterization with multimodal imaging. Retina. 2010;30(9):1441–1454.

12. Spaide RF, Curcio CA, Zweifel SA. Drusen, an old but new frontier. Retina. 2010;30(8):1163–1165.

13. Spaide RF. Improving the age-related macular degeneration construct: A new classification system. Retina. 2018;38(5):891–899.

14. Fleckenstein M, Keenan TDL, Guymer RH, et al. Age-related macular degeneration. Nature Reviews Disease Primers. 2021;7(1):31.

15. Sarks J, Arnold J, Ho IV, Sarks S, Killingsworth M. Evolution of reticular pseudodrusen. Br J Ophthalmol. 2011;95(7):979–985.

16. Anderson DMG, Kotnala A, Migas LG, et al. Lysolipids are prominent in subretinal drusenoid deposits, a high-risk phenotype in age-related macular degeneration. Front Ophthalmol (Lausanne). 2023;3.

17. Spaide RF. Outer retinal atrophy after regression of subretinal drusenoid deposits as a newly recognized form of late age-related macular degeneration. Retina. 2013;33(9):1800–1808.

18. Suzuki M, Sato T, Spaide RF. Pseudodrusen subtypes as delineated by multimodal imaging of the fundus. Am J Ophthalmol. 2014;157(5):1005–1012.

19. Won J, Yaghy A, Ploner SB, et al. High-resolution, motion-corrected, volume-fused oct for investigating longitudinal changes in subretinal drusenoid deposits in intermediate amd. Transl Vis Sci Technol. 2025;14(6):15.

20. Wu Z, Ayton LN, Makeyeva G, Guymer RH, Luu CD. Impact of reticular pseudodrusen on microperimetry and multifocal electroretinography in intermediate age-related macular degeneration. Invest Ophthalmol Vis Sci. 2015;56(3):2100–2106.

21. Zhang Y, Sadda SR, Sarraf D, et al. Spatial dissociation of subretinal drusenoid deposits and impaired scotopic and mesopic sensitivity in amd. Invest Ophthalmol Vis Sci. 2022;63(2):32.

22. Sassmannshausen M, Dongelci S, Vaisband M, et al. Spatially resolved association of structural biomarkers on retinal function in non-exudative age-related macular degeneration over 4 years. Invest Ophthalmol Vis Sci. 2024;65(4):45.

23. Sassmannshausen M, Pfau M, Thiele S, et al. Longitudinal analysis of structural and functional changes in presence of reticular pseudodrusen associated with age-related macular degeneration. Invest Ophthalmol Vis Sci. 2020;61(10):19.

24. Goerdt L, Amjad M, Swain TA, et al. Extent and topography of subretinal drusenoid deposits associate with rod-mediated vision in aging and amd: Alstar2 baseline. Invest Ophthalmol Vis Sci. 2024;65(10):25.

25. Sivaprasad S, Bird A, Nitiahpapand R, et al. Perspectives on reticular pseudodrusen in age-related macular degeneration. Surv Ophthalmol. 2016;61(5):521–537.

26. Domalpally A, Agrón E, Pak JW, et al. Prevalence, risk, and genetic association of reticular pseudodrusen in age-related macular degeneration: Age-related eye disease study 2 report 21. Ophthalmology. 2019;126(12):1659–1666.

27. Agron E, Domalpally A, Cukras CA, et al. Reticular pseudodrusen: The third macular risk feature for progression to late age-related macular degeneration: Age-related eye disease study 2 report 30. Ophthalmology. 2022;129(10):1107–1119.

28. Agron E, Domalpally A, Chen Q, et al. An updated simplified severity scale for age-related macular degeneration incorporating reticular pseudodrusen: Age-related eye disease study report number 42. Ophthalmology. 2024;131(10):1164–1174.

29. Sadda SR, Guymer R, Holz FG, et al. Consensus definition for atrophy associated with age-related macular degeneration on oct: Classification of atrophy report 3. Ophthalmology. 2018;125(4):537–548.

30. Burns SA, Elsner AE, Sapoznik KA, Warner RL, Gast TJ. Adaptive optics imaging of the human retina. Prog Retin Eye Res. 2019;68:1–30.

31. Williams DR, Burns SA, Miller DT, Roorda A. Evolution of adaptive optics retinal imaging [invited]. Biomed Opt Express. 2023;14(3):1307–1338.

32. Zhang Y, Wang X, Rivero EB, et al. Photoreceptor perturbation around subretinal drusenoid deposits as revealed by adaptive optics scanning laser ophthalmoscopy. Am J Ophthalmol. 2014;158(3):584–596 e581.

33. Zhang Y, Wang X, Godara P, et al. Dynamism of dot subretinal drusenoid deposits in age-related macular degeneration demonstrated with adaptive optics imaging. Retina. 2018;38(1):29–38.

34. Zhang Y, Wang X, Sadda SR, et al. Lifecycles of individual subretinal drusenoid deposits and evolution of outer retinal atrophy in age-related macular degeneration. Ophthalmology Retina. 2020;4(3):274–283.

35. Paavo M, Lee W, Merriam J, et al. Intraretinal correlates of reticular pseudodrusen revealed by autofluorescence and en face oct. Invest Ophthalmol Vis Sci. 2017;58(11):4769–4777.

36. Chen L, Messinger JD, Kar D, Duncan JL, Curcio CA. Biometrics, impact, and significance of basal linear deposit and subretinal drusenoid deposit in age-related macular degeneration. Invest Ophthalmol Vis Sci. 2021;62(1):33.

37. Querques G, Canoui-Poitrine F, Coscas F, et al. Analysis of progression of reticular pseudodrusen by spectral domain-optical coherence tomography. Invest Ophthalmol Vis Sci. 2012;53(3):1264–1270.

38. Jaffe GJ, Chakravarthy U, Freund KB, et al. Imaging features associated with progression to geographic atrophy in age-related macular degeneration: Classification of atrophy meeting report 5. Ophthalmology Retina. 2021;5(9):855–867.

39. Zhang Y, Wang X, Clark ME, Curcio CA, Owsley C. Imaging of age-related macular degeneration by adaptive optics scanning laser ophthalmoscopy in eyes with aged lenses or intraocular lenses. Transl Vis Sci Technol. 2020;9(8):41.

40. Xu X, Liu X, Wang X, et al. Retinal pigment epithelium degeneration associated with subretinal drusenoid deposits in age-related macular degeneration. Am J Ophthalmol. 2017;175:87–98.

41. Xu X, Wang X, Sadda SR, Zhang Y. Subtype-differentiated impacts of subretinal drusenoid deposits on photoreceptors revealed by adaptive optics scanning laser ophthalmoscopy. Graefes Arch Clin Exp Ophthalmol. 2020;258(9):1931–1940.

42. Wang X, Sadda SR, Ip MS, Sarraf D, Zhang Y. In vivo longitudinal measurement of cone photoreceptor density in intermediate age-related macular degeneration. Am J Ophthalmol. 2023;248:60–75.

43. Davis MD, Gangnon RE, Lee LY, et al. The age-related eye disease study severity scale for age-related macular degeneration: Areds report no. 17. Arch Ophthalmol. 2005;123(11):1484–1498.

44. Meadway A, Girkin CA, Zhang Y. A dual-modal retinal imaging system with adaptive optics. Opt Express. 2013;21(24):29792–29807.

45. Meadway A, Wang X, Curcio CA, Zhang Y. Microstructure of subretinal drusenoid deposits revealed by adaptive optics imaging. Biomed Opt Express. 2014;5(3):713–727.

46. Yu Y, Zhang Y. Dual-thread parallel control strategy for ophthalmic adaptive optics. Chinese optics letters : COL. 2014;12(12):121202.

47. Yu Y, Zhang T, Meadway A, Wang X, Zhang Y. High-speed adaptive optics for imaging of the living human eye. Opt Express. 2015;23(18):23035–23052.

48. Sevilla MB, McGwin G, Jr., Lad EM, et al. Relating retinal morphology and function in aging and early to intermediate age-related macular degeneration subjects. Am J Ophthalmol. 2016;165:65–77.

49. Spaide RF, Curcio CA. Drusen characterization with multimodal imaging. RETINA. 2010;30(9):1441–1454.

50. Balaratnasingam C, Cherepanoff S, Dolz-Marco R, et al. Cuticular drusen: Clinical phenotypes and natural history defined using multimodal imaging. Ophthalmology. 2018;125(1):100–118.

51. Voichanski S, Bousquet E, Abraham N, et al. En face oct and the phenotype characterization of drusen. Invest Ophthalmol Vis Sci. 2025;66(9):52.

52. Voichanski S, Bousquet E, Abraham N, et al. En face optical coherence tomography illustrates the trizonal distribution of drusen and subretinal drusenoid deposits in the macula. Am J Ophthalmol. 2024;261:187–198.

